# High risk of SARS-CoV-2 infection among frontline healthcare workers in Northeast Brazil: a respondent-driven sampling approach

**DOI:** 10.1101/2021.10.08.21264755

**Authors:** Maria de Fátima Pessoa Militão de Albuquerque, Wayner Vieira de Souza, Ulisses Ramos Montarroyos, Cresio Romeu Pereira, Cynthia Braga, Thália Velho Barreto de Araújo, Ricardo Arraes de Alencar Ximenes, Demócrito de Barros Miranda-Filho, Célia Landmann Szwarcwald, Paulo Roberto Borges de Souza Junior, Morgana do Nascimento Xavier, Clarice Neuenschwander Lins de Morais, Gabriela Diniz Militao de Albuquerque, Cristiane C. Bresani Salvi, Carolline de Araújo Mariz, Noêmia Teixeira de Siqueira Filha, Jadson Mendonça Galindo, Cláudio Luiz de França Neto, Jessyka Mary Vasconcelos Barbosa, Maria Amelia de Sousa Mascena Veras, Luana Nepomuceno Gondim Costa Lima, Luciane Nascimento Cruz, Carl Kendall, Ligia Regina Franco Sansigolo Kerr, Celina Maria Turchi Martelli

## Abstract

**Introduction:** The disparities in the risk of severe acute respiratory syndrome coronavirus 2 (SARS-CoV-2) infection among frontline health care workers (HCWs) and the unique work circumstances are poorly documented for low-and middle-income countries.

**Methods:** We assessed the frequency of SARS-CoV-2 infection, personal protective equipment (PPE) shortages, PPE use, and accidents involving biological material among HCWs in the Recife metropolitan area, Northeast Brazil. Using respondent driven sampling, we included HCWs attending suspected or confirmed COVID-19 patients from May 2020 to February 2021.

**Results:** We analyzed 1,525 HCWs (527 physicians, 471 registered nurses, 263 nursing assistants/technicians, and 264 physical therapists). Women predominated in all categories (81.1%). Nurses were older and had more comorbidities (hypertension and overweight/obesity) than the other HCWs. The overall prevalence of SARS-CoV-2 infection was 61.8% after adjustment for the cluster random effect, weighted by network, and reference population size. The independent risk factors for a positive RT-PCR test were being a nursing assistant (OR adjusted: 2.56), not always using all recommended PPE in routine practice (ORadj: 2.15), and reporting a splash of biological fluid/respiratory secretion in the eyes (ORadj: 3.37).

**Conclusions:** The high risk of infection among HCWs reflects PPE shortages and younger, possibly less experienced, frontline HCWs. There were disparities in the risk of SARS-CoV-2 infection among HCWs, with nursing assistants being the most vulnerable, possibly due to their longer and frequent contact with COVID-19 patients.

## Introduction

The unprecedented rapid spread of severe acute respiratory syndrome coronavirus 2 (SARS-CoV-2) and its potentially severe outcomes have highly impacted the healthcare system, the global economy, and security.^1,2^ According to the World Health Organization (WHO), the global cumulative number of confirmed coronavirus disease 2019 (COVID-19) cases had reached approximately 190.5 million with four million deaths by July 19, 2021.^3^ In Brazil, approximately 19 million COVID-19 cases and 514,000 related deaths were reported within the same period. These figures represent almost 10% and 13% of the global COVID-19 cases and registered deaths, respectively, yet the Brazilian population represents approximately 2.5% of the global population. Since the beginning of the pandemic, the federal government has opposed the recommendations for social distancing and individual protection measures while endorsing ineffective pharmaceutical interventions, hampering the epidemic control efforts of the public health authorities at the state and municipal levels.^4^

Healthcare workers (HCWs) are considered a high-risk group due to the nature of their work. An Anglo-American prospective cohort that included approximately 100,000 HCWs showed a 3.4-fold higher risk of COVID-19 among frontline workers compared with the general community.^5^ This comprehensive study used an online survey with the advantage of potentially avoiding personal contact during the pandemic, as well as allowing timely responses and dissemination of results.^5^ A systematic review and meta-analysis, covering the period from the inception of the pandemic to July 2020, included 46 studies: approximately 70% were conducted in Europe (n=31), nine in the USA, six in Asia, and none in Latin America. Among symptomatic HCWs, the pooled overall prevalence of SARS-CoV-2 infection was 19% using reverse transcription-polymerase chain reaction (RT-PCR).^6^

In the Americas, 569,304 COVID-19 cases, including 2,506 deaths, had been reported among HCWs by August 2020.^7^ According to public health surveillance, approximately 32% of Mexico City HCWs (n=11,226) had been infected with SARS-CoV-2 by July 2020.^8^ Additionally, cross-sectional studies conducted in Brazil, Colombia, and Ecuador revealed lack of personal protective equipment (PPE) among 70% of frontline workers in the early pandemic response.^9^ In Brazil, studies conducted using RT-PCR in teaching hospitals showed a varying prevalence of SARS-CoV-2 infection (42.4%–15%).^10,11,12^ However, information on the prevalence of SARS-CoV-2 infection among frontline HCWs and risk factors for most regions of Brazil is limited.

This study assessed the prevalence of SARS-CoV-2 infection and evaluated PPE shortages, use of individual protective measures, and biological accidents among HCWs in Recife metropolitan area of Northeast Brazil.

## Methods

### Study design

This prospective study assessed the frequency of infected HCWs and their risk factors, using the respondent-driven sampling (RDS) methodology, and collecting data with a smartphone-based application. RDS was chosen as a sampling approach for two main reasons: restrictions in conducting face-to-face interviews due to lockdown and the lack of a frame list of frontline HCWs attending emergency rooms, hospitals, and new field hospitals. RDS approach is based upon direct participant involvement.

The baseline findings are described following the Strengthening the Reporting of Observational Studies in Epidemiology (STROBE) guidelines for RDS.^13^

### Setting

The study was conducted in the Recife metropolitan region, Pernambuco State, Northeast Brazil, where the first COVID-19 case was reported on March 12, 2020 The peak of the pandemic was during the 21^st^ epidemiologic week in 2020.^14,15^ This densely populated region comprises 15 municipalities with approximately four million inhabitants, corresponding to 42% of the state population.^16^ The Brazilian unified health system (Sistema Unico de Saude— SUS) has provided universal coverage since 1990, with heterogeneity among the regions.^17^

### Formative research

Formative research (FR) was conducted with the four HCW categories included in the study (physicians, nurses, nurse assistants, and physical therapists). The FR applied in-depth interviews to explore workplace changes, use and access to PPE, routine attendance, and possible acceptability of the study.

### Participants

We recruited HCWs attending suspected or confirmed COVID-19 patients from May 21, 2020 to February 10, 2021. Recruitment started with five “seeds” for each category, non-randomly selected from the target population. We asked each participant to identify five other members of the same professional network category, providing their names and mobile phone numbers to the fieldworkers. The process continued until a suitable sample size was reached. This study did not offer any incentive.

We calculated a sample size of 1,100 HCWs, considering a 95% confidence level (CI) to estimate a 40% prevalence of infections with a 5% error and a design effect of three.

The network size of each HCW was measured by the final answer to the following questions:

1) “How many colleagues do you know, who also know you by name, work in the Recife metropolitan region and are assisting COVID-19 patientsã”, 2) “How many of those colleagues have been in professional contact with you in the last two weeksã,” and 3) “How many of them are close to you and you would invite to participate in this studyã.”

### Variables

The variables collected were adapted from the WHO interim guidance (March 2020) on health workers’ exposure risk assessment and management in the context of the COVID-19 pandemic. The variables were:

1. Age, sex, and professional category;
2. Self-reported comorbidities (diabetes mellitus, hypertension, overweight or obesity, cardiopathy, nephropathy, and others);
3. Healthcare attending—public or private sector, outpatient, emergency rooms and intensive care units (ICU); number of healthcare facilities.
4. Adherence to infection prevention and control (IPC). We checked for gloves, medical masks, face shields, goggles or protective glasses, and waterproof aprons.
5. Adherence to IPC when performing aerosol-generating procedures (AGPs) using the abovementioned grading criteria. In this section, we added the N95 respirator. The variables related to adherence to IPC (items 4 and 5) were grouped as always versus not always.
6. Accidents with biological material—I) during the period of healthcare interaction and II) if there was an accident with biological fluid or respiratory secretions, which type it was (splash in the mucous membrane of eyes, mouth, or nose; non-intact skin; and puncture-sharp accident).^18^ The outcome was a self-reported positive RT-PCR test for SARS-CoV-2.

### Data collection

Data were collected using a web-based software platform by FITec (Recife, Pernambuco, Brazil). The HCWs answered the questionnaire by accessing a link that could be opened on a smartphone or a computer browser. Providing electronic informed consent was mandatory to participate and access the questionnaire. The project was approved by the National Ethics Committee (CONEP; CAAE: 30629220.8.0000.0008).

### Data analysis

Participants were weighted by the size of each category, provided by each professional board, and by the inverse of the size of their professional network, based on the following question: “How many of these colleagues are close to you and would you invite to participate in this studyã” To avoid the influence of extreme network sizes on the weight of each professional, we limited the network size to 3 to 150 for outlier correction.^19^ For missing data—representing around 8% of the total—we used available information from the other two questions related to network size, and when necessary, we applied the overall mean of the stratum. We considered the recruitment of participants by the same professional as an RDS cluster.

Categorical variables are presented as percentages and 95% CIs by HCW category and overall frequencies adjusted for the design. The chi-squared test was used for comparison between groups. We calculated the means, medians, and 95% CIs for continuous variables. Bivariate analysis was performed to assess the association between potential risk factors and RT-PCR positivity. Variables associated with the outcome at *p*<0.20 were included in the multivariate model. In the final model, we considered variables at the *p*<0.10 level statistically significant. All statistical analyses were performed using Stata, version 15.0 (StataCorp LLC, College Station, TX, USA).

## Results

### Participants

Of the 2,474 participants recruited, 1,525 (527 physicians, 471 registered nurses, 263 nursing assistants, and 264 physical therapists) completed the questionnaire (Figure1). Figure 2 illustrates the recruitment chain for each category.

**Figure 1.**
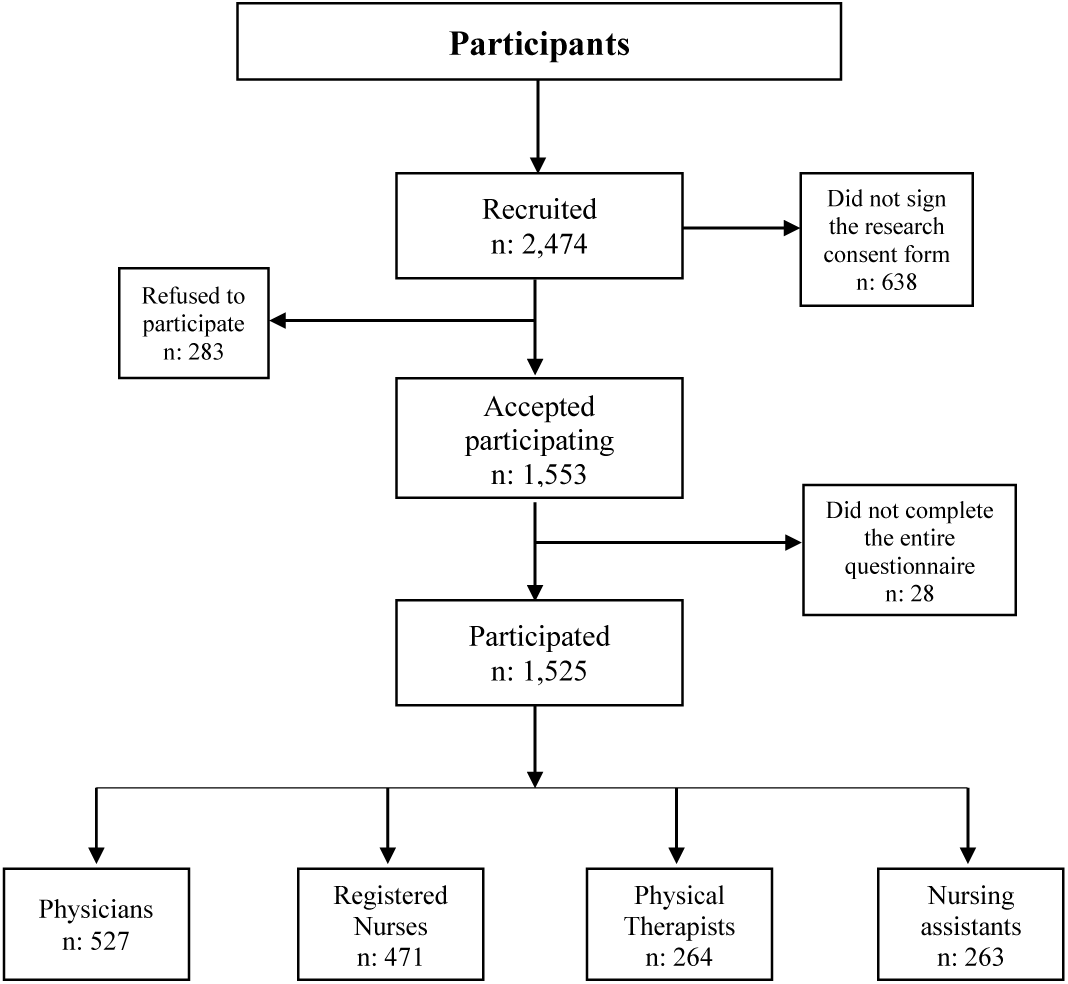
Participant flowchart.

**Figure 2.**
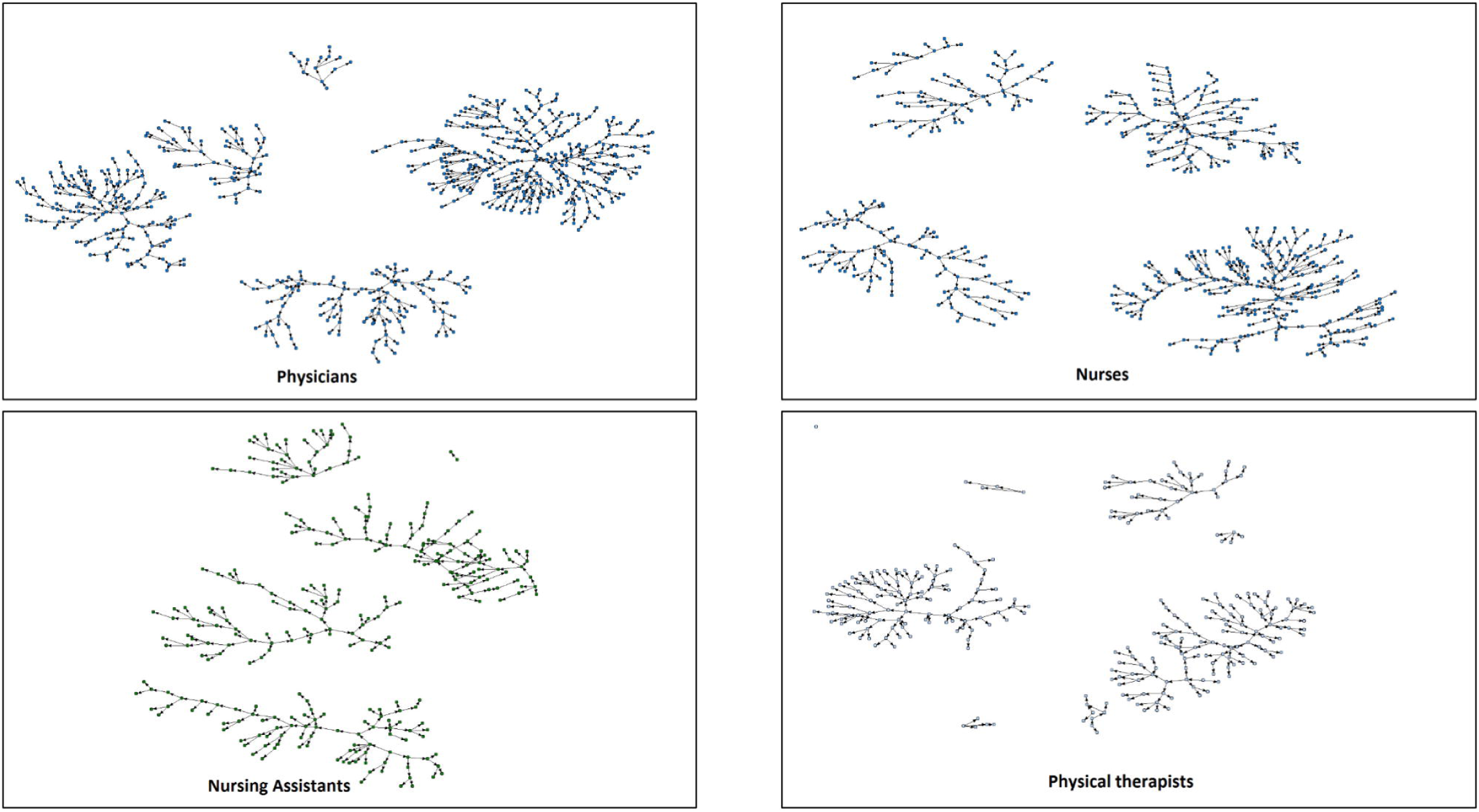
Respondent-driven sampling recruitment chains.

### Descriptive data

Overall, women represented 81.1% of the sample after adjustment to the reference population and for the study design (Table 1). Women also predominated in all professional categories, with the lowest percentage among physicians (63.4%) and the highest among nurses (86.7%) and nursing assistants (85.5%). The age distribution was as follows: 32.7% and 35.6% were <30 and 30–39 years old, respectively. Only 0.1% of the participants were aged ≥60 years. Physicians and physical therapists were the youngest groups, comprising 56.6% and 45.0%, respectively, of those 20–29 years old. Comorbidities affected 30.0% of the studied population. Overweight/obesity (12.6%) and hypertension (11.9%) were the most prevalent comorbidities among nursing assistants and nurses than among the other categories. In total, 71.4% of HCWs attended COVID-19 cases exclusively in the public sector, including hospitals, emergency units, ambulance services, and primary care units. Most HCWs (73.5%) worked either in emergency rooms or ICU. Notably, 55.8% of the physicians and 37.8% of the physical therapists indicated working in three or more institutions during the pandemic (Table 1). Overall, 78.0% of the participants received training on the use of PPE. Physical therapists (87.0%) and nursing assistants (81.1%) received a higher and similar frequency of training compared to the other categories. Almost half of the HCWs (47.7%) reported a shortage of PPE items during the COVID-19 pandemic. Regarding wearing PPE in routine activities, the overall frequencies varied widely for each item: 90.1% for single-use gloves to 29.9% for face shields. Most HCWs (82.2%) reported performing AGPs on COVID-19 patients. Almost all participants reported having always used single-use gloves (98.4%) and N95 respirators (86.4%) during AGPs. The N95/PPF2 respirator was reused for more than seven days by approximately 28.3% of the participants, with highest and lowest frequencies reported by physicians (49.3%) and nursing assistants (20.6%), respectively. Overall, 63.7% of the HCWs reported always wearing all PPE items as recommended by the WHO. The self-perception of SARS-CoV-2 risk of infection in the previous 15 days varied: 33.4% for “performing a procedure on a patient with COVID-19;” 17.7% for “sharing the break room with their colleagues;” 16% for the “reuse of N95 respirators;” 10.6% for the “use of poor quality PPE;” 10.2% during “doffing;” 9.6% for “working with colleagues with COVID-19 symptoms;” 1.9% for “lack of PPE in the service;” and 0.5% for “donning PPE.” HCWs reported 186 episodes of exposure to biological fluids/respiratory secretions during healthcare interaction with COVID-19 patients. Accidents were more frequent among physicians (13.9%) and less frequent among physical therapists (7.6%) (Table 2).

**Table 1.**
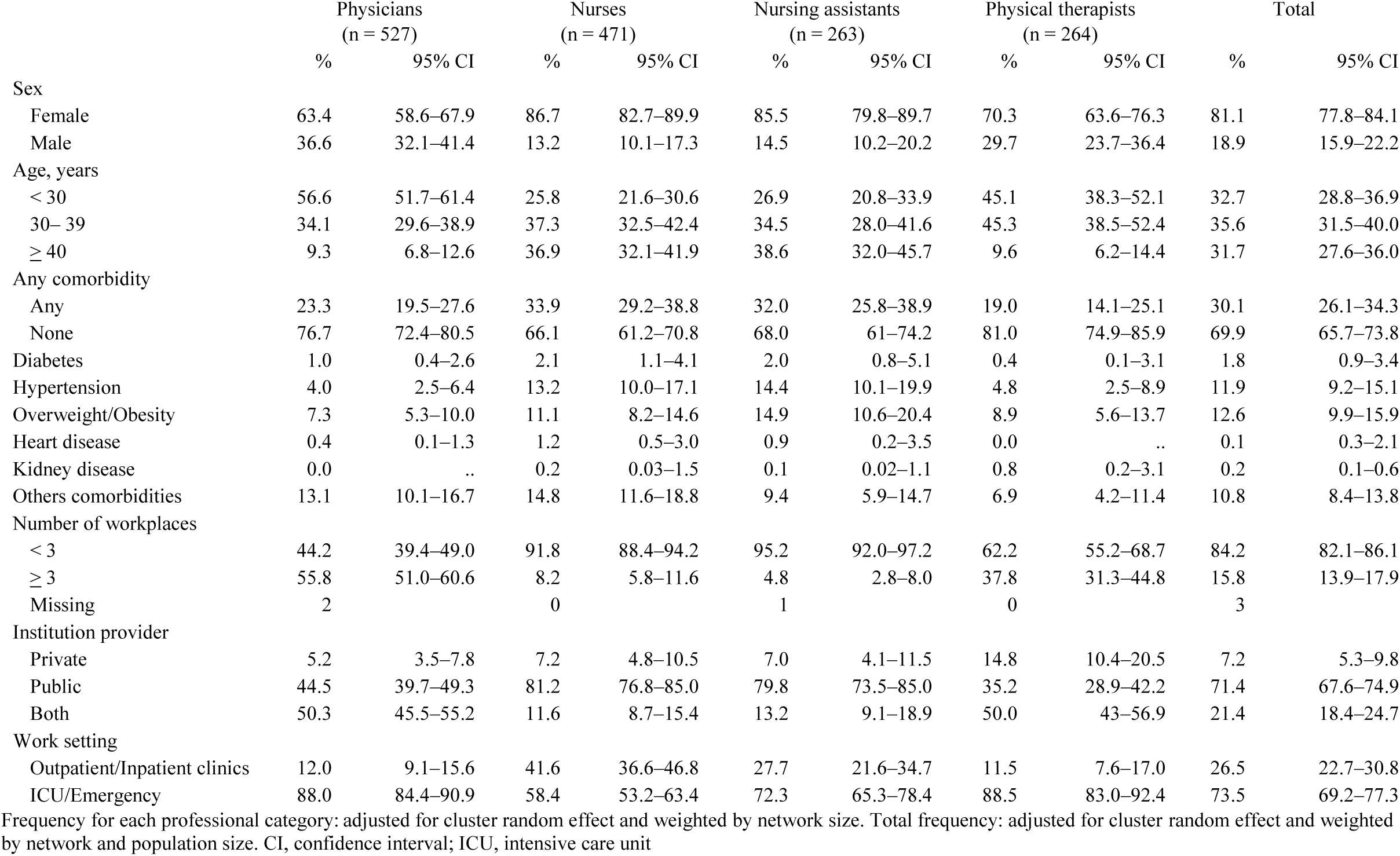
Demographic, clinical, and working baseline characteristics of health care workers in the metropolitan region of Recife, Northeast Brazil, 2020 to 2021.

**Table 2.**
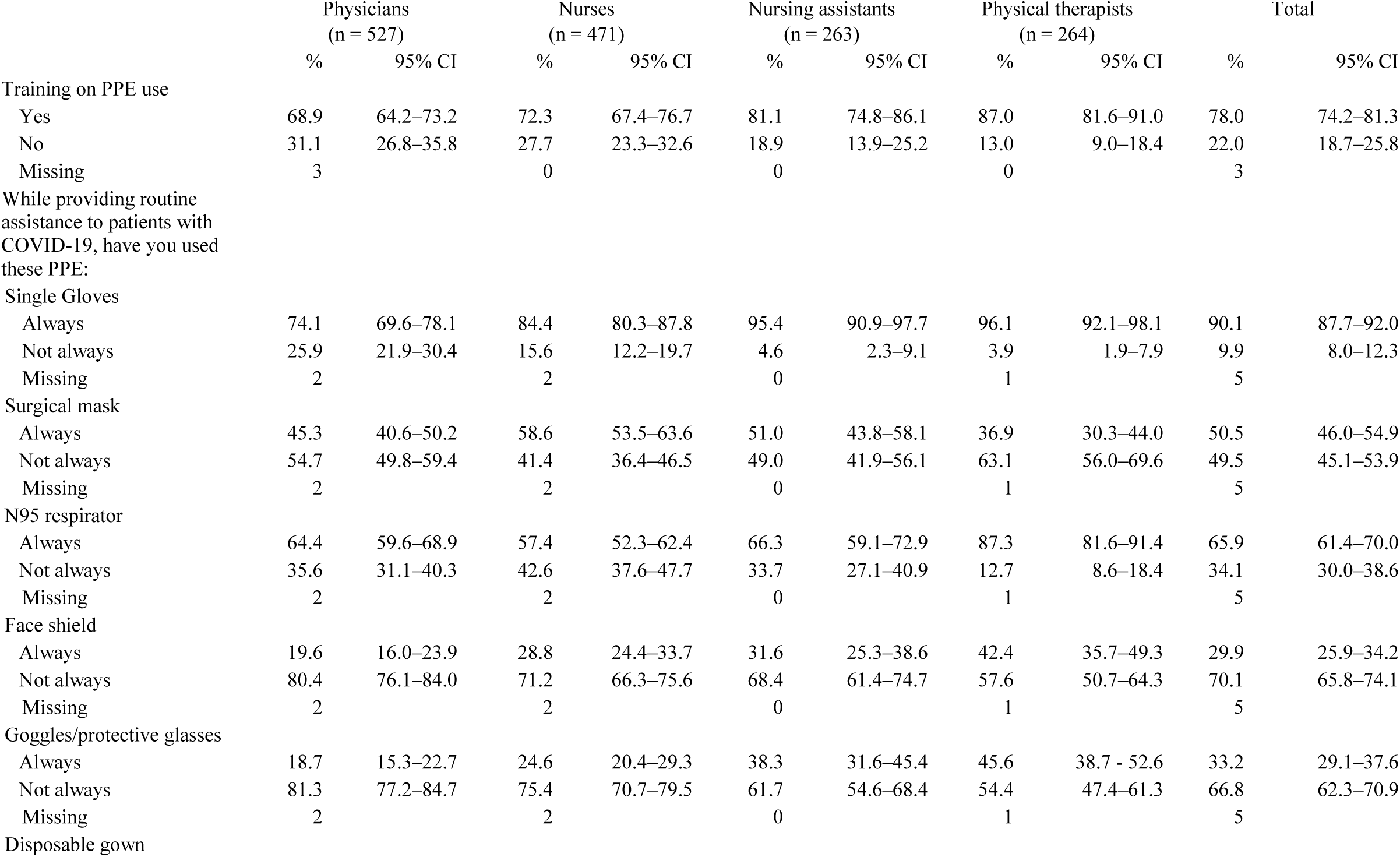
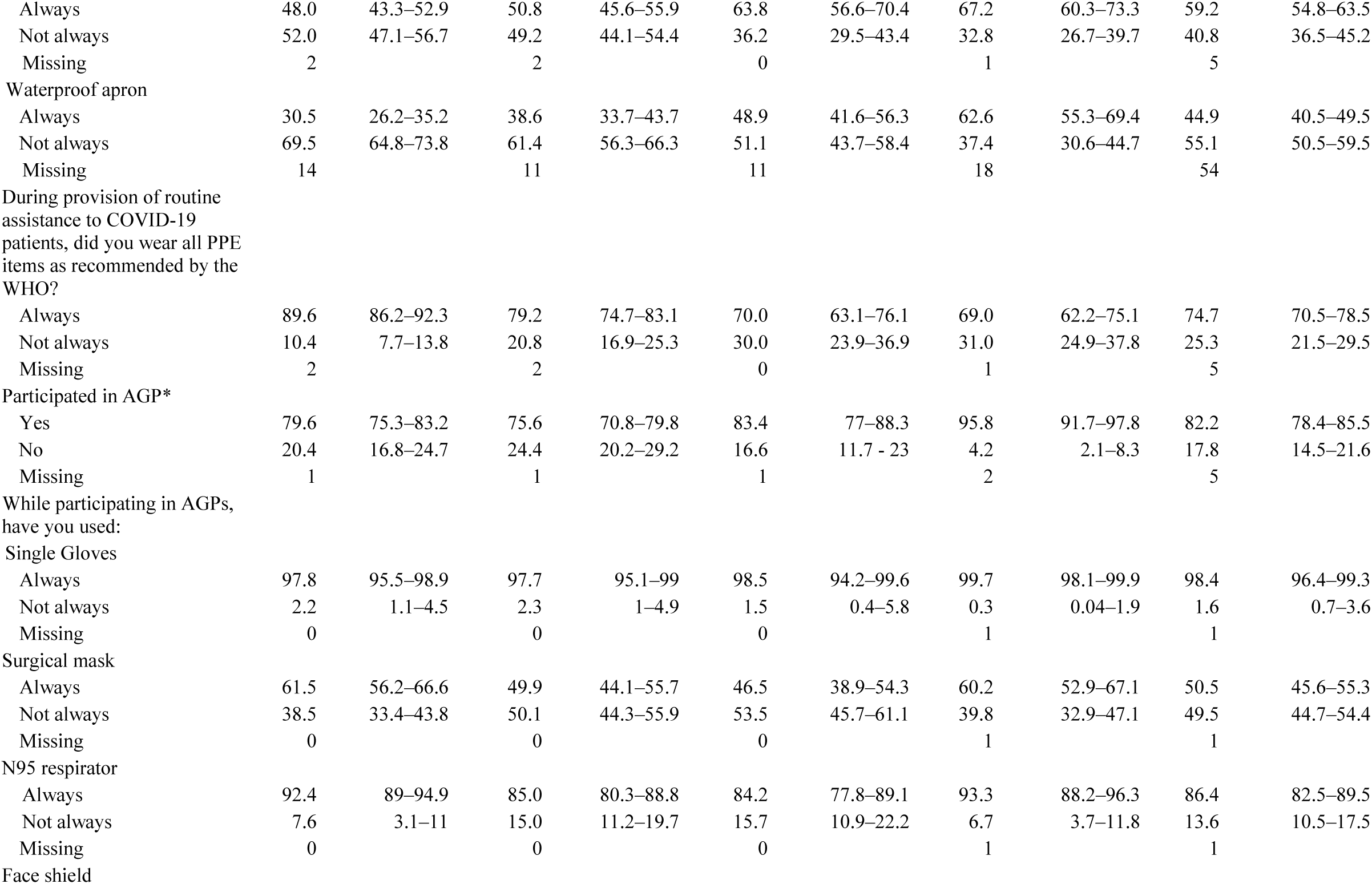
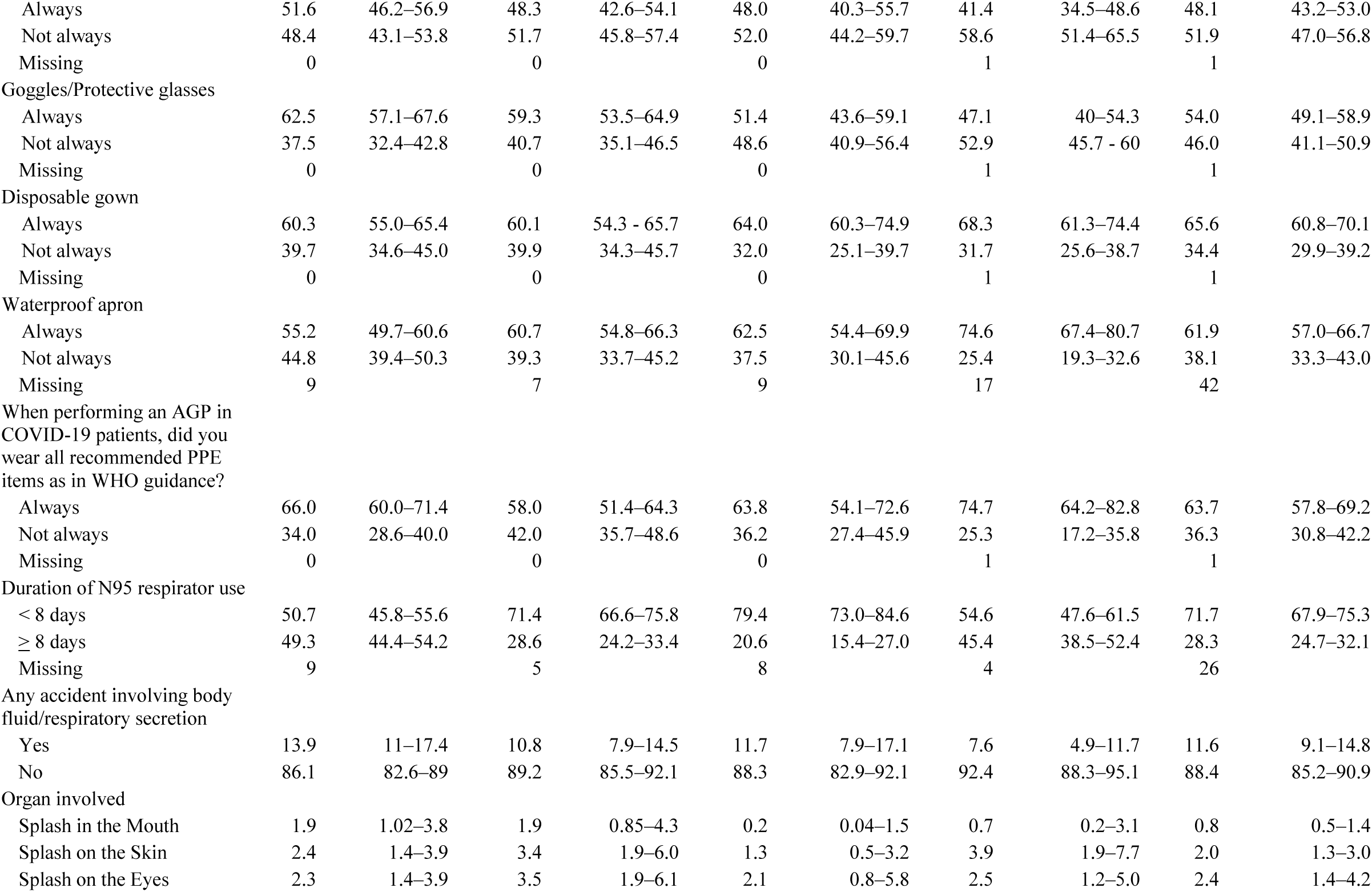
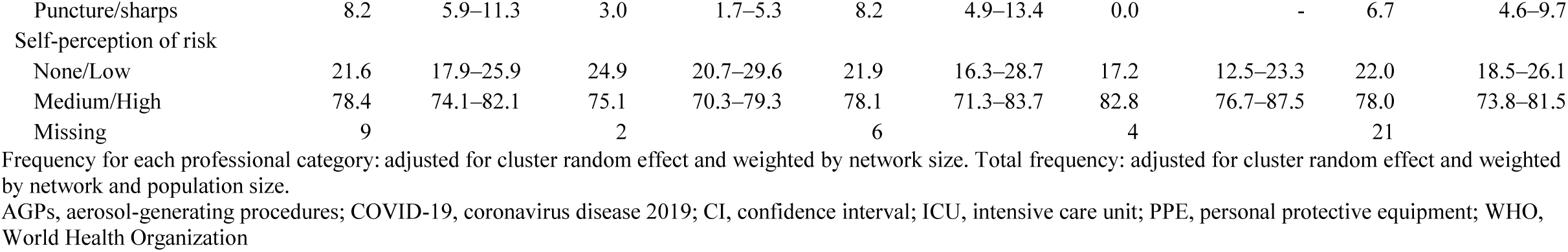
Adherence to infection prevention and control during healthcare interactions with COVID-19 patients and accidents with biological materials.

The frequency of COVID-19 testing varied from 41.2% for physical therapists to 51.1% for physicians. Individuals with any comorbidity were more likely to get tested (56.8%) than those without comorbidities (*p*<0.001). HCWs who worked in three or more health services were also more likely to get tested (54.9%) than those who worked in only one health service (42.1%) (*p*<0.001). There was no statistical difference in the likelihood of testing, according to sex, age group (<30 versus ≥30 years old), work setting (outpatients, inpatients, and emergency rooms and ICU), self-perception of risk (no risk to high risk of exposure), reported accidents with biological fluid/respiratory secretion, and when performing AGPs (Supplementary Table 1). For the tested HCWs, mostly symptomatic, the overall self-reported SARS-CoV-2 infection was 61.8% after adjustment for random cluster effects, weighted by network and population size. The highest infection positivity was among nursing assistants (70.0%), followed by physicians (55.0%), physical therapists (54.7%), and nurses (48.1%), adjusted for random cluster effects (Figure 3). RT-PCR screening was performed mainly among symptomatic cases in all categories, ranging from 81.8% to 91.8% for physicians and nursing assistants, respectively.

**Figure 3.**
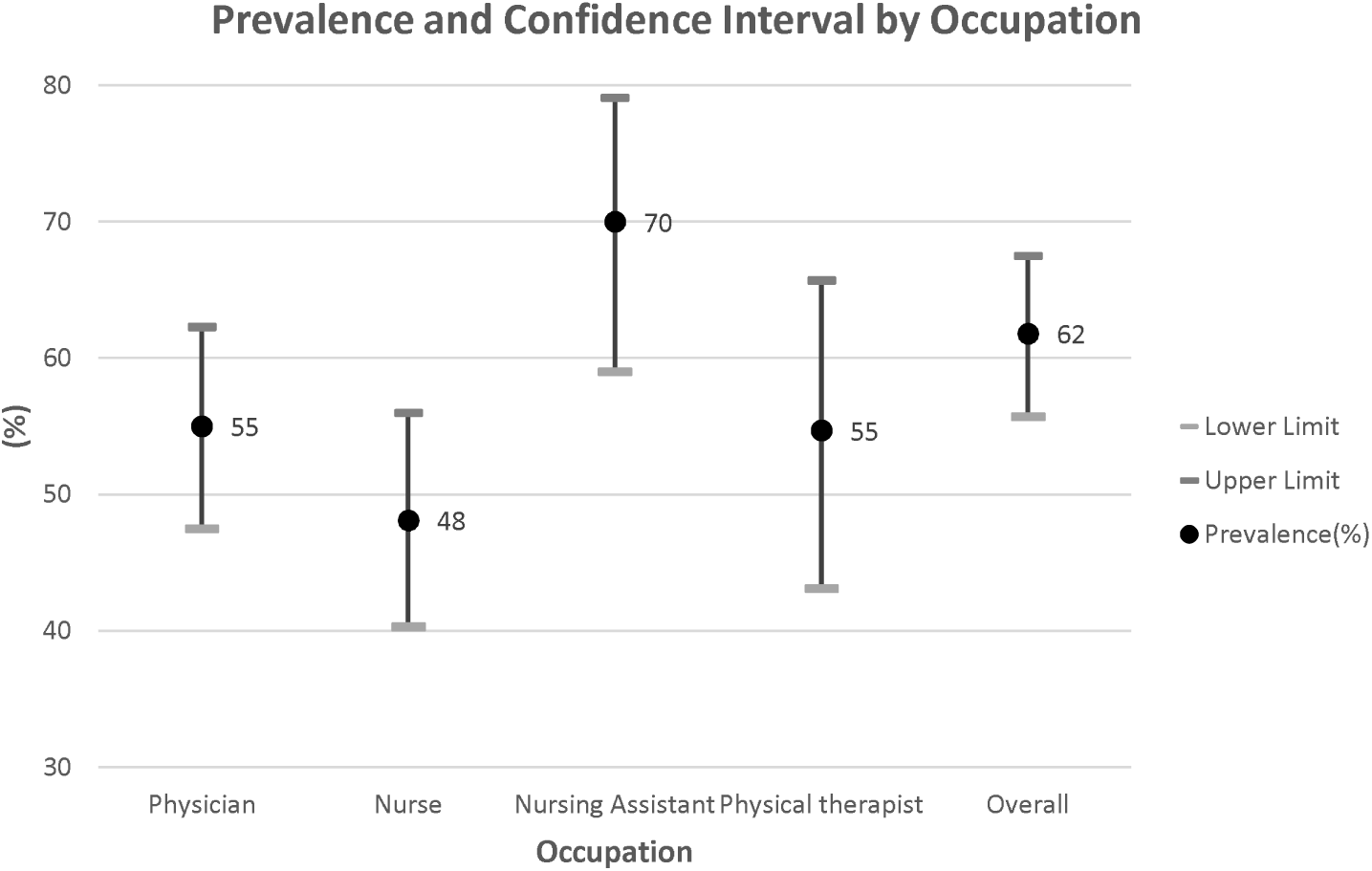
Frequencies of self-reported SARS-CoV-2 infection by healthcare categories.

Almost half of the HCWs (47.8%) reported taking sick leave due to COVID-19, with a similar trend among the other categories (*p*=0.159). The median length of health leave was 14 days for all professional categories, reflecting a standard procedure. Of 399 symptomatic SARS-CoV-2 infected HCWs, 10% (n=41) were hospitalized.

In a bivariate analysis, the nursing assistant category was positively associated with infection (odds ratio [OR]=2.77, *p*<0.001) compared to nurses. Reporting any accident involving body fluid/respiratory secretion was associated with infection (OR=2.67, *p*<0.014). When considering each accident, splashes in the eyes were a stronger predictor of infection (OR=4.07, *p*<0.031). During routine assistance of COVID-19 patients, not always wearing the complete set of recommended PPE items was associated with infection (OR=2.14; *p*=0.013) when compared to always using PPE. Not always using the complete recommended PPE items during AGPs was also associated with infection (OR=1.69; *p*=0.063) when compared with always using PPE (Table 3).

**Table 3.** Potential risk factors for reporting a positive PCR COVID-19 result among front line healthcare professionals.

|  | Odds Ratio | 95% CI | P-value |
| --- | --- | --- | --- |
| Sex |  |  |  |
| Female | 1.0 | .. | .. |
| Male | 1.35 | 0.78–2.34 | 0.288 |
| Age, years | 1.03 | 0.65–1.64 | 0.889 |
| Occupation |  |  |  |
| Nurse | 1.0 | .. | .. |
| Physical therapist | 1.42 | 0.88–2.27 | 0.148 |
| Physician | 1.32 | 0.91–1.91 | 0.142 |
| Nursing Assistant | 2.77 | 1.64–4.67 | <0.001 |
| Any comorbidity | 1.19 | 0.75–1.90 | 0.454 |
| Number of workplaces |  |  |  |
| < 3 | 1.0 | .. | .. |
| ≥ 3 | 0.83 | 0.53–1.30 | 0.428 |
| Institution provider |  |  |  |
| Private | 1.0 | .. | .. |
| Public | 0.92 | 0.42–2.02 | 0.844 |
| Both | 0.93 | 0.41–2.10 | 0.863 |
| Work setting |  |  |  |
| Outpatient /Inpatient clinics | 1.0 | .. | .. |
| ICU/Emergency | 1.54 | 0.92–2.60 | 0.102 |
| Training on PPE use | 1.06 | 0.62–1.80 | 0.829 |
| Any accident involving body fluid/respiratory secretion | 2.67 | 1.22–5.82 | 0.014 |
| Splash in the mouth |  |  |  |
| No accident | 1.0 | .. | .. |
| Yes | 3.84 | 0.64–22.95 | 0.140 |
| Other accident | 2.30 | 0.85–6.23 | 0.102 |
| Splash on the skin |  |  |  |
| No accident | 1.0 | .. | .. |
| Yes | 1.86 | 0.54–6.44 | 0.328 |
| Other accident | 2.50 | 0.80–7.85 | 0.116 |
| Splash in the eyes |  |  |  |
| No accident | 1.0 | .. | .. |
| Yes | 4.07 | 1.14–14.55 | 0.031 |
| Other accident | 2.07 | 0.71–6.08 | 0.184 |
| Puncture/sharp accident |  |  |  |
| No accident | 1.0 | .. | .. |
| Yes | 2.25 | 0.51–9.89 | 0.282 |
| Other accident | 2.51 | 1.10–5.72 | 0.028 |
| Duration N95 respirator use |  |  |  |
| < 8 days | .. | .. | .. |
| ≥ 8 days | 0.96 | 0.59–1.55 | 0.869 |
| Used All PPE items during AGP# |  |  |  |
| Did not Always use | 1.68 | 0.97–2.92 | 0.063 |
| Used all PPE items while assisting COVID-19 patients |  |  |  |
| Yes | 1.0 | .. | .. |
| No | 2.14 | 1.18–3.88 | 0.013 |
| Time on the front-line, days | 0.997 | 0.994–1.000 | 0.042 |

Adjusted for cluster random effect and weighted by network and population size. AGP, aerosol-generating procedure; COVID-19, coronavirus disease 2019; CI, confidence interval; ICU, intensive care unit; PPE, personal protective equipment

In the final multivariate logistic regression model, the following were risk factors for infection: being a nursing assistant (OR adjusted=2.56, *p*=0.002), not always having used PPE during care of patients with COVID-19 (OR adjusted=2.15, *p*=0.044), and having suffered a splash to the eyes (OR adjusted=3.37, *p*=0.034) (Table 4).

**Table 4.** Final multivariate model for factors associated with reported positive PCR COVID-19 results.

|  | Odds Ratio | 95% CI | *P*-value |
| --- | --- | --- | --- |
| Occupation |  |  |  |
| Nurse | 1.0 | .. | .. |
| Physical therapist | 1.47 | 0.80–2.72 | 0.214 |
| Physician | 1.20 | 0.76–1.90 | 0.426 |
| Nursing assistant | 2.56 | 1.42–4.61 | 0.002 |
| Splash on the eyes |  |  |  |
| No accident | 1.0 | .. | .. |
| Yes | 3.37 | 1.10–10.34 | 0.034 |
| Any accident | 1.59 | 0.51–4.90 | 0.421 |
| Used all PPE items while assisting patients with COVID-19 |  |  |  |
| Yes | 1.0 | .. | .. |
| No | 2.15 | 1.02–4.53 | 0.044 |

Adjusted for cluster random effect and weighted by network and population size
COVID-19, coronavirus disease 2019; CI, confidence interval; PPE, personal protective equipment

## Discussion

The current study showed substantial heterogeneity in demographic and self-referred comorbidities between HCW categories during the COVID-19 pandemic. Of note, physicians and physical therapists at the frontline were younger and mainly worked in the ICU and emergency rooms when compared with nurses. This reflects the expansion of the healthcare workforce with the inclusion of younger physicians and physical therapists, possibly inexperienced professionals, forcibly driven to work as front liners in a high-risk environment. Nurses and nursing assistants were older and reported more comorbidities, particularly hypertension and overweight/obesity. According to the accumulated evidence, the public health strategy was to prevent exposure among older age groups and/or individuals with comorbidities, as older age and comoridities are strong prognostic factors for hospitalization and death.^20^

To our knowledge, our study depicted one of the highest frequencies of SARS-CoV-2 infections among HCWs. One likely explanation is that most of the participants tested were symptomatic, reflecting the policy of making RT-PCR tests for COVID-19 diagnosis available to frontline HCWs. Thus far, there has been no mass RT-PCR testing strategy for the Brazilian population despite WHO recommendations.^21^ Worldwide, the prevalence closest to that of our study was 55%, by RT-PCR among 177 symptomatic medical residents in New York City at the beginning of the COVID-19 pandemic.^22^ In Southeast Brazil, a high prevalence of SARS-CoV-2 infection (42%) tested by RT-PCR was found among symptomatic HCWs at a teaching hospital in Sao Paulo, from March to May 2020.^10^ Another study found a prevalence of 14% (701 out of 4,987) using RT-PCR in a group composed of mainly symptomatic HCWs, at a hospital in the south of Brazil from April to June 2020.^12^ This variation might be attributable to the dynamics of the pandemic in different regions of the country, the availability/quality of PPE, and training in different healthcare settings.

Our study found a 7% prevalence of infection (by RT-PCR) among the 105 asymptomatic HCWs, which is similar to the overall 5% prevalence of infection found by a large screening study for SARS-CoV-2 infection in the metropolitan area of Mexico City.^23^ As expected, these results reflect the positive predictive value of clinical manifestations. Although seroprevalence studies cannot be directly compared to our findings, the frequencies of SARS-CoV-2 infection among HCWs in São Paulo city ranged from 5.5% (IgG ELISA) in a private hospital to 14% (IgG/IgM antibody, WONDFO™) in a large public hospital in 2020.^11,24^ Both hospital settings stated that they adopted high-quality hospital infection control and provided complete PPE in the early stages of the COVID-19 pandemic. This may reflect especially high-quality healthcare facilities in more developed regions of the country and the rates reported were similar to those reported in another meta-analysis of seroprevalence studies.^25^

Critical aspects for the high risk of SARS-CoV-2 infection included shortage of PPE items reported by approximately half the HCWs. Moreover, 22% of HCWs reported not been trained on PPE use. The lack of preparedness of the health workforce to respond to the COVID-19 pandemic was not only encountered by low- and medium-income countries like Brazil but also in high-income countries at the beginning of the pandemic.^26^ At the individual level, one-fourth of the HCWs reported that PPE was not always used according to the WHO recommendations.^21^ When performing AGPs, the nursing staff had the highest frequency (over 35%) of not fully adhering to complete PPE.^27^ Furthermore, not using the recommended PPE during routine attendance of COVID-19 cases caused a 2.2-fold increased risk of a SARS-CoV-2 positive RT-PCR test result. Accidents with biological fluids occurred in all categories, however, they were most frequently reported among physicians, the youngest, and perhaps the group with the least experience working in critical conditions. Reporting an accident with biological fluids, such as a splash in the eye, was positively associated with infection in the final multivariable model. Although it is uncertain whether viruses occasionally present in biofluids are infectious, these fluids should be considered potentially infectious.^28^ Moreover, the eye has been considered a possible route of SARS-CoV-2 entry through drainage via the nasolacrimal duct to the upper respiratory tract.^29^ These accidents with biological fluids should be further investigated in other studies, as recommended by the WHO guidelines.^18^ The prevalence among HCWs in the current study was at least 20-fold higher when compared to the 3.2% seroprevalence in a population-based survey using SARS-CoV-2 antibody rapid tests conducted during the first wave of the pandemic in the same region.^30^ Therefore, there is strong evidence that HCWs are at a high risk of SARS-CoV-2 infection in low- and medium-income settings, such as Northeast Brazil.

To the best of our knowledge, this is the largest Latin American study of HCWs during the COVID-19 pandemic, with the inclusion of the four main healthcare professionals in the public and private sectors and multiple levels of health services. Previous investigations conducted in Brazil were mainly restricted to one hospital setting and did not apply the WHO questionnaire.^18^ One advantage of using the RDS methodology was that it allowed the inclusion of frontline HCWs from different healthcare settings, including the private and public health services, providing a more comprehensive picture of frontline HCWs during the pandemic. Furthermore, as HCWs worked in more than one health service and/or in newly implemented “field hospitals/units,” this strategy allowed us to capture the full extent of characteristics of the workforce and the risk factors for infection. Another advantage of applying an online questionnaire was to avoid face-to-face interviews during the lockdown and/or social distancing restrictions, reduce errors in data transcription, and obtain timely results.

This study had some limitations. First, there was an imbalance in recruitment among the HCW categories; physicians and nurses were more rapidly enrolled by RDS than nursing assistants. One possible explanation is that physicians and nurses seem to understand research methodology better and/or to have either better smartphones or data plans required to answer the approximately 15-minute online questionnaire. Physicians and nurses were also a more vocal category early in the pandemic, publicizing the constraints/pressure of the workplace. Conversely, nursing assistants, as routine healthcare assistants, spend more time providing direct patient care and have low wages. They could also be less confident/willing to participate due to work overload or unfavorable socio-economic conditions when compared to the other categories that require university degrees. Additionally, disclosure of the work environment concerning PPE and infection control prevention may be problematic for nursing assistants whose jobs are less stable and more prone to replacement in our setting. The current study did not discriminate the source of SARS-CoV-2 infection among HCWs. Accidents involving biological fluids should be further investigated in other studies to validate this finding.

Finally, our findings provide a comprehensive picture of the factors associated with SARS-CoV-2 infection among HCWs. This study highlighted the high prevalence of SARS-CoV-2 infection among all HCW categories, with nursing assistants being the most affected.

## Data Availability

Proposals for the dataset (de-identified participant data, data dictionary) should be directed to the corresponding author. To gain access, data requestors will need to present their plan of analysis and sign a data access agreement.

## Ethics statements

Providing electronic informed consent was mandatory to participate and access the questionnaire. The project was approved by the National Ethics Committee (CONEP; CAAE: 30629220.8.0000.0008).

## Acknowledgments

We thank HCWs for their participation. We acknowledge the Institute of Health Technology Assessment and MCTIC/CNPq/FNDCT/MS/SCTIE/Decit Nº 07/2020 for support. The following researchers received scholarship (CNPq-Pq): 308974/2018-2 to CMTM, 309722/2017-9 to RAAX, 301905/2017-7 to MFPMA, no. 30735/2018-1 to CB and 303661/2017-8 to WVS. CRP received scholarship from CNPq, EV-1 no. 315877/2020-0.

## Author contributions

MFPMA, WVS, CMTM, RAAX, DBMF, TB, CK, and LRFSK contributed to the study concept and design. CB, MNX, CNLM, GDMA, CBS, CAM, NTSF, JMG, CLFN, and JMVB contributed to the acquisition of data. MFPMA, URM, WVS, CLS, PRBSJ, and CRP contributed to the data analysis and creation of tables and figures. MFPMA, WVS, CMTM, RAAX, DMF, TVBA, MASMV, LNGCL, CB, and LNC contributed to data interpretation. MFPMA, WVS, URM have verified the underlying data. CMTM, MFPMA, WVS, and CRP drafted the initial manuscript and all other coauthors contributed scientific inputs equally towards the interpretation of the findings and the final draft of the manuscript. All authors confirm that they had full access to all the data in the study and accept responsibility to submit for publication.

