## Supplementary Table 1 for "High risk of SARS-CoV-2 infection among frontline healthcare workers in Northeast Brazil: a respondent-driven sampling approach"

**Supplementary Table 1. Characteristics of the study population according to RT-PCR testing**

| | | RT-PCR testing | | *P*-value |
|---|---|---|---|---|
| | | **Yes (%)** | **No (%)** | |
| **Occupation category** | | | | 0.02 |
| | Physician | 269 (51.1) | 257 (48.9) | |
| | Registered nurse | 224(47.6) | 247(52.4) | |
| | Nursing assistant | 110 (42.0) | 152 (58.0) | |
| | Physical therapist | 108(41.2) | 154(58.8) | |
| **Sex** | | | | 0.43 |
| | Female | 530 (46.2) | 618 (53.8) | |
| | Male | 181 (48.5) | 192 (51.5) | |
| **Age group, years** | | | | 0.15 |
| | < 30 | 523 (45.7) | 622 (54.3) | |
| | ≥ 30 | 188 (50.0) | 188 (50.0) | |
| **Any comorbidity** | | | | < 0.001 |
| | Yes | 246 (56.8) | 187 (43.2) | |
| | No | 465 (42.7) | 623 (57.3) | |
| **Number of workplaces (hospitals/clinics)** | | | | < 0.01 |
| | <3 | 247 (54.0) | 210 (46.0) | |
| | ≥3 | 462 (43.5) | 599 (56.5) | |
| **Work setting** | | | | 0.39 |
| | Emerg/ICU | 565 (47.3) | 629 (52.7) | |
| | Outpat/Inpatients | 146 (44.7) | 181 (55.3) | |
| **Institution provider** | | | | < 0.001 |
| | Private | 48 (42.1) | 66 (57.9) | |
| | Public | 393 (43.0) | 522 (57.0) | |
| | Both | 270 (54.9) | 222 (45.1) | |
| **Performed aerossol** | | | | 0.36 |
| **generating procedure** | | | | |
| | Yes | 600 (47.3) | 669 (52.7) | |
| | No | 110 (44.5) | 137 (55.5) | |
| | Missing | 1 (20.0) | 4 (80.0) | |
| **Same N95 respirator, use duration, days** | | | | 0.023 |
| | ≤ 7 | 458 (49.00 | 476 (51.0) | |
| | > 7 | 243 (43.0) | 322 (57.0) | |
| **Self-perceived risk** | | | | 0.85 |
| | None/Low | 36 (45.1) | 43 (54.9) | |
| | Medium/High | 665 (46.7) | 760 (53.3) | |
| **Accident involving biological fluid/respiratory secretion** | | | | 0.644 |
| | Yes | 84 (45.2) | 102 (54.8) | |
| | No | 627 (47.0) | 708 (53.0) | |
| **Sick leave due to** | | | | < 0.001 |
| **COVID-19 symptoms** | | | | |
| | Yes | 576 (79.7) | 147 (20.3) | |
| | No | 130 (16.5) | 659 (83.5) | |
| **Had COVID-19-like symptoms/signs** | | | | < 0.001 |
| | Yes | 601 (68.2) | 280 (31.8) | |
| | No | 110 (17.0) | 530 (82.8) | |

COVID-19, coronavirus disease 2019; RT-PCR, reverse transcription polymerase chain reaction
